# Understanding the impact of COVID-19 risk perceptions on mitigation behaviors: A mixed methods approach using survey instruments and serious games

**DOI:** 10.1101/2022.08.07.22278512

**Authors:** Scott C. Merrill, Sarah A. Nowak, Trisha R. Shrum, John P. Hanley, Eric M. Clark, Luke Fredrickson, Tung-Lin Liu, Robert M. Beattie, Aislinn O’Keefe, Asim Zia, Christopher J. Koliba

## Abstract

COVID-19 risk mitigation behavior, including social distancing and mask wearing, was a principal factor influencing the spread of COVID-19. Yet this behavior, and its association with COVID-19 perceptions and beliefs, is poorly understood. Here we used a mixed methods approach combining serious game data with survey instruments to describe relationships between perceptions and behavior. Using a series of survey questions, participants were described along a spectrum denoting their perception of their susceptibility to COVID-19 associated with a list of activities. Afterwards, participants engaged with a serious game to examine behavioral responses to scenarios involving shopping at a grocery store and going to a park during simulated pandemic conditions. Messages describing the simulated infection risk were shown to drive many behavioral decisions. Another significant correlate, derived from survey results, was the participant’s perception of susceptibility associated with various activities for acquiring the COVID-19 infection. Individuals that perceived every day activities, such as grocery shopping, as unlikely to lead to a COVID-19 infection spent more time near others in the game-simulated grocery store environment compared to those that consider such activities as risky. Additionally, we found that participant behavior became increasingly risky as time progresses if they were lucky enough not to experience an infection. This reflects behavior observed in the United States and more broadly, possibly explains how people update their perception of the risk of activities. Overall, results show a link between perception and action with regards to COVID-19 and support the use of targeted risk messaging to influence behavior. Moreover, the link between reported real-world perceptions and game behavior suggest that serious games can be used as valuable tools to test policies, risk messaging and communication, with the goal of nudging individuals with varied and nuanced perceptions and belief sets towards behaviors that will reduce the impact of COVID-19.

## Introduction

Appropriate use of non-pharmaceutical risk mitigating behaviors is crucial for controlling the spread of infectious diseases such as COVID-19. In the United States, individual use of risk mitigating behaviors, such as social distancing and mask wearing has been inconsistent (1, 2). Drivers of risk mitigating behaviors include social components (3-5) observable, in part, through communities that share attitudes and beliefs (6). A variety of public health policies and interventions have been instituted to increase the use of risk mitigating behaviors such as obtaining vaccines (7). However, these policies have been designed and enacted with a limited understanding of the underpinnings of behavioral decisions, leading to a critical need to examine motivators of risk mitigation behaviors. The United States government’s “Operation Warp Speed” was designed to develop vaccines as quickly as possible but neglected social and behavioral aspects, such as messaging that would explain and encourage vaccine use (8). This serves as a contemporary example of why assumptions of ubiquitous responses and rational behavior are flawed and can be detrimental. Yet, examining risk behavior is challenging because motivating factors are variable and complex (9).

Understanding how we make decisions is paramount to creating systems where more optimal or resilient solutions can be reached. Yet studying the vagaries of human behavior and decision-making is challenging because of the wide variety of rational and irrational behavior, and the difficulties in collecting behavioral data. While traditional methods, such as surveys, generate critical data, they may fail to capture unbiased responses (10), especially in risk management situations where survey participants are distanced from real-world scenarios (11-13) (14). Moreover, the lack of dynamism inherent in these methods limits insights into essential drivers of decision-making. I.e., if a hypothetical situation evolves or respondents learn or adapt over time, these behavioral effects may not be adequately captured with hypothetical survey questions. An alternative to traditional methods, and an under-utilized method for gathering data, is through the use of serious games designed primarily for data collection. For decades, economic and social game-theory style experiments, such as Flood and Dresher’s (15) Prisoner’s Dilemma, or economic experiments such as Ellsburg’s (16) ambiguity aversion experiments or Holt and Laury’s (17) risk aversion studies have been used to assess behavior and decision-making. It has been found that participants respond dynamically to observed changes during the experiment (e.g., experiencing a conflict), and thus provide information about how individuals process information and make decisions as situations evolve and/or individual perceptions are modified by their experiences. Moreover, serious games provide the opportunity to use performance-based incentives which have been found to increase engagement and the salience of decisions, and further align the incentives of participants with the underlying research structure and (18-20). Serious game data can be integrated with other traditional data streams to get a nuanced, realistic representation of behavioral responses. Thus a robust approach to gaining a fuller understanding of human nature may be through triangulating a variety of methods, such as surveys, focus groups, as well as serious games, then to integrate data streams into data analyses.

Computer-based serious games allow users to “experience” hypothetical scenarios without placing them in real decision-making situations, thus allowing for imposed conflict to incite a behavioral response. Serious gaming research has been applied to simulated disease outbreaks in livestock, and has exposed sharply irrational and boundedly rational behavior when participants were confronted with conflict and exposed to dynamic risk situations (21-26). For example, willingness to invest in protective measures for the health of livestock was influenced by treatments that provided or withheld certain types of information (22, 23). Interestingly, the type of information withheld prompted different situational responses (23). Specifically, withholding information about disease in the system reduced willingness to invest in biosecurity measures, while withholding information about how others were responding to the disease outbreak increased willingness to invest in biosecurity. Merrill et al. (24) found that in some risk scenarios, participants complied with suggested rules three-times more frequently when presented with risk information with a visual threat gauge compared to when risk was communicated using a number. Interactive, immersive or virtual reality imagery has been shown to alter intent more than passive information transfer (e.g., reading) (27). Graphical or visual displays have been identified as increasing salience in certain contexts over simple text or numerical displays (19). One’s local environment and imagery are known to influence affect (28), which in turn is known to influence decision-making (29).

One theoretical framework that suggests a mostly rational framework for decision-making processes associated with health behaviors is the Health Belief Model (HBM). In the HBM, perceptions of preventive behaviors and disease influence behavioral intention and behaviors (30). One of the key HBM tenets is that perceived susceptibility influences mitigation behavior, specifically where those that perceive themselves as less susceptible, are less likely to adopt behaviors that reduce the threat (31, 32). We suggest that collecting data on beliefs of the safety of COVID-19 related activities and observations of social distancing behaviors will provide evidence for decision-making heuristics that can be further used to help design policies and incentives to help reduce the impact of COVID-19 (33). Further, experimental treatments that vary the situational context could be used to examine COVID-19 risk perceptions, and thus gather data to assess communication strategies, the results of which could be used to increase mitigation behaviors.

We used survey methodologies triangulated with serious games to collect human belief, perception and behavioral data. We used survey questions to understand perceived risk of activities, then we used a serious game to study risk mitigating behavior using treatments that vary both direct and indirect COVID-19 infection risk information (Fig. 1). Drawing from a sample population from across the United States, we sought to understand relationships between COVID-19 risk perceptions and the effect on behavior. We looked for behavioral differences when participants were confronted with a series of simulated everyday activities, i.e., obtaining food and getting exercise. We hypothesized that perceptions of the risk or safety associated with many everyday activities would be associated with risk behavior in simulated activities. Testing this hypothesis will help us understand linkages between risk perceptions, intent and consequent behavior, as well as how risk messages can influence behavior. Further, results will provide insight into how to effectively sculpt messages and policies to nudge behavior towards compliance with COVID-19 risk mitigating behavior.

**Figure 1.**
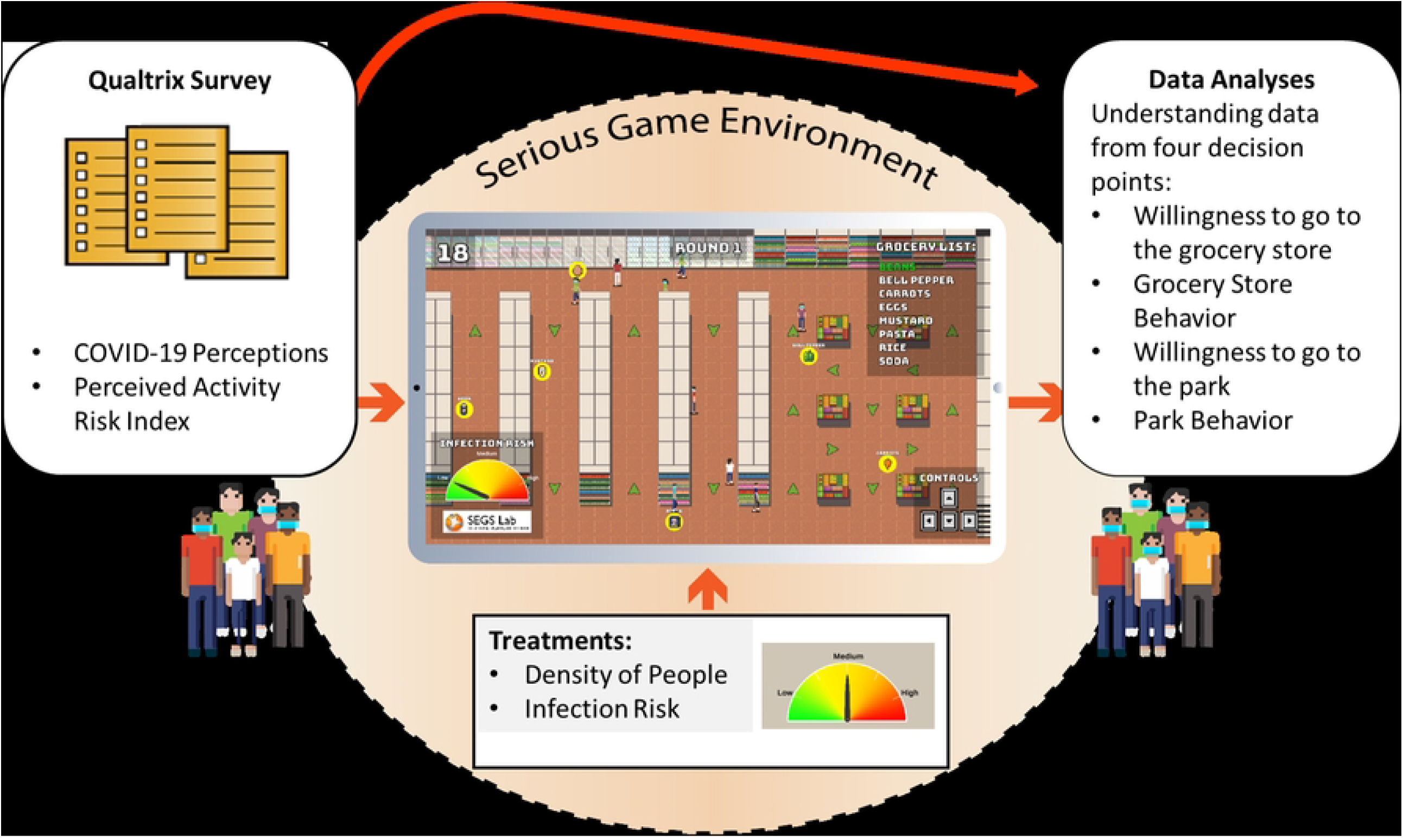
Project design including a Qualtrics survey linked to the serious game, followed by data analysis.

## Methods

### Recruitment of Participants

Five hundred and sixty-six (566) individuals were recruited and participated in the experiment between March 9^th^, 2021 and April 14^th^, 2021 (before widespread availability of vaccines). Individuals were recruited through the online workplace Amazon Mechanical Turk (21, 34, 35), an online recruitment tool that has been found to be largely as representative as simple convenience samples (36). Recruits were informed that they would be paid based on their performance during the experiment (i.e., performance-based incentives). Participants started the experiment by completing a survey on the Qualtrics platform before following an online link to initiate the serious game. The serious game portion started with an interactive demonstration to frame the game mechanics and decision mechanism. Institutional Review Board protocols were followed for an experiment using human participants (University of Vermont IRB # CHRBSS-15-319-IRB). On average, participants earned $10.14 with a minimum payout of $7.94 and a maximum of $12.33.

### Survey Details

Participants started the experiment by completing a survey which was implemented using Qualtrics software (survey results are also analyzed in a separate paper (37)). For the present analysis, we constructed a Perceived Activity Risk Index from seventeen survey questions related to the perception of the safety of different activities in regards to exposure to COVID-19 (Table 1). For example, one of the questions from the set asked “How safe or unsafe are the following actions for avoiding exposure to coronavirus: Obtaining take-out food from a restaurant.”? (1: Extremely Safe, 2: Somewhat Safe, 3: Unsure, 4: Somewhat Unsafe, 5: Extremely Unsafe). The Perceived Activity Risk Index was designed to quantify the perceived safety by summing the response values from the seventeen questions with summed values ranging from a possible low of 17 (all activities deemed to be Extremely Safe) to 85 (all activities deemed to be Extremely Unsafe). The summed values were then normalized by subtracting the minimum and dividing by the range. Resultant values thus varied from 0-1 with 0 equating to perceiving activities as being extremely safe and 1 being those that perceived activities as being extremely unsafe. Those individuals falling on the lower end of this scale may believe that COVID-19 was not a real or serious threat. The Perceived Activity Risk Index was considered the measure of the participant’s perceived susceptibility, thus aligning the risk index with the perception of susceptibility tenet of the Health Belief Model.

**Table 1.**
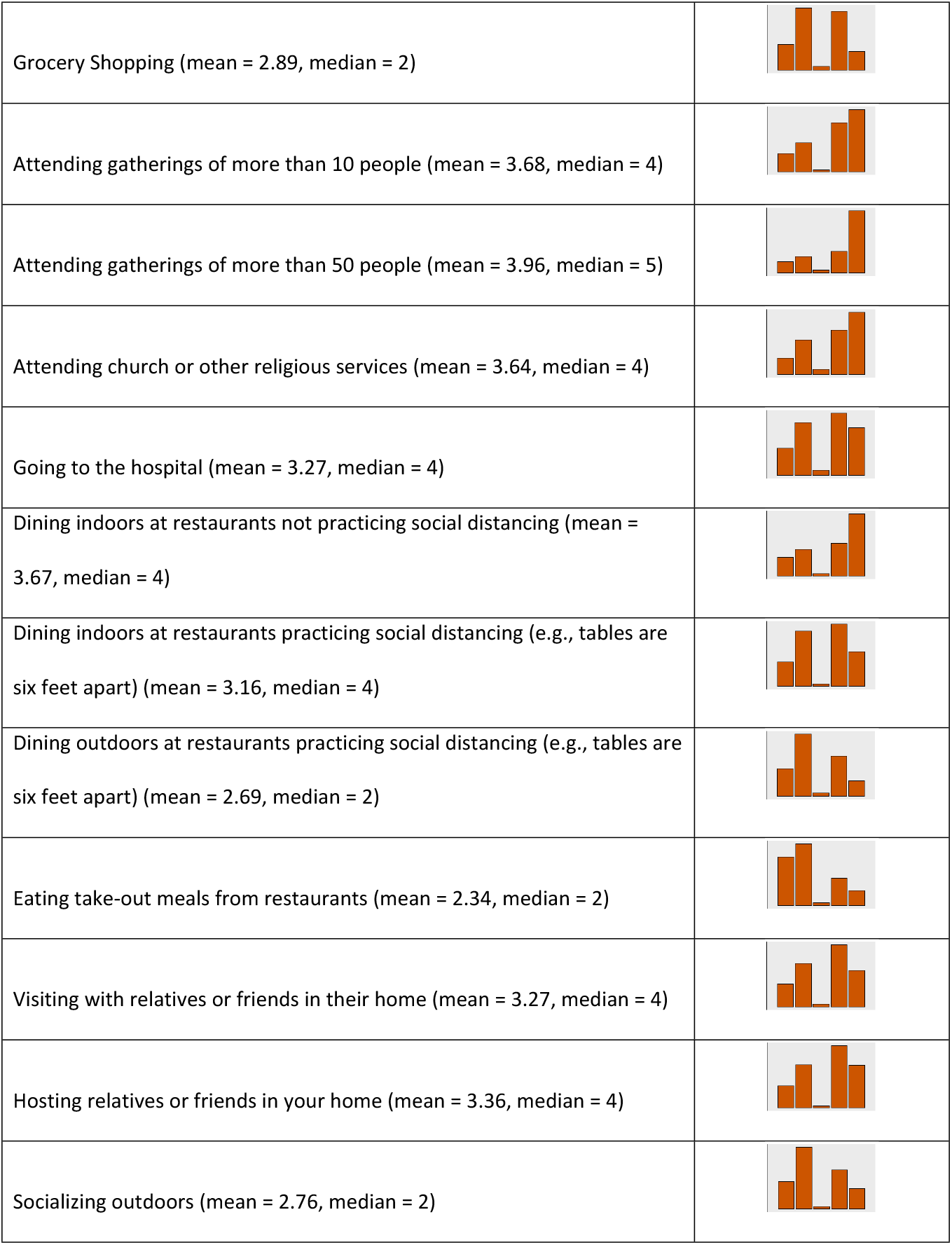
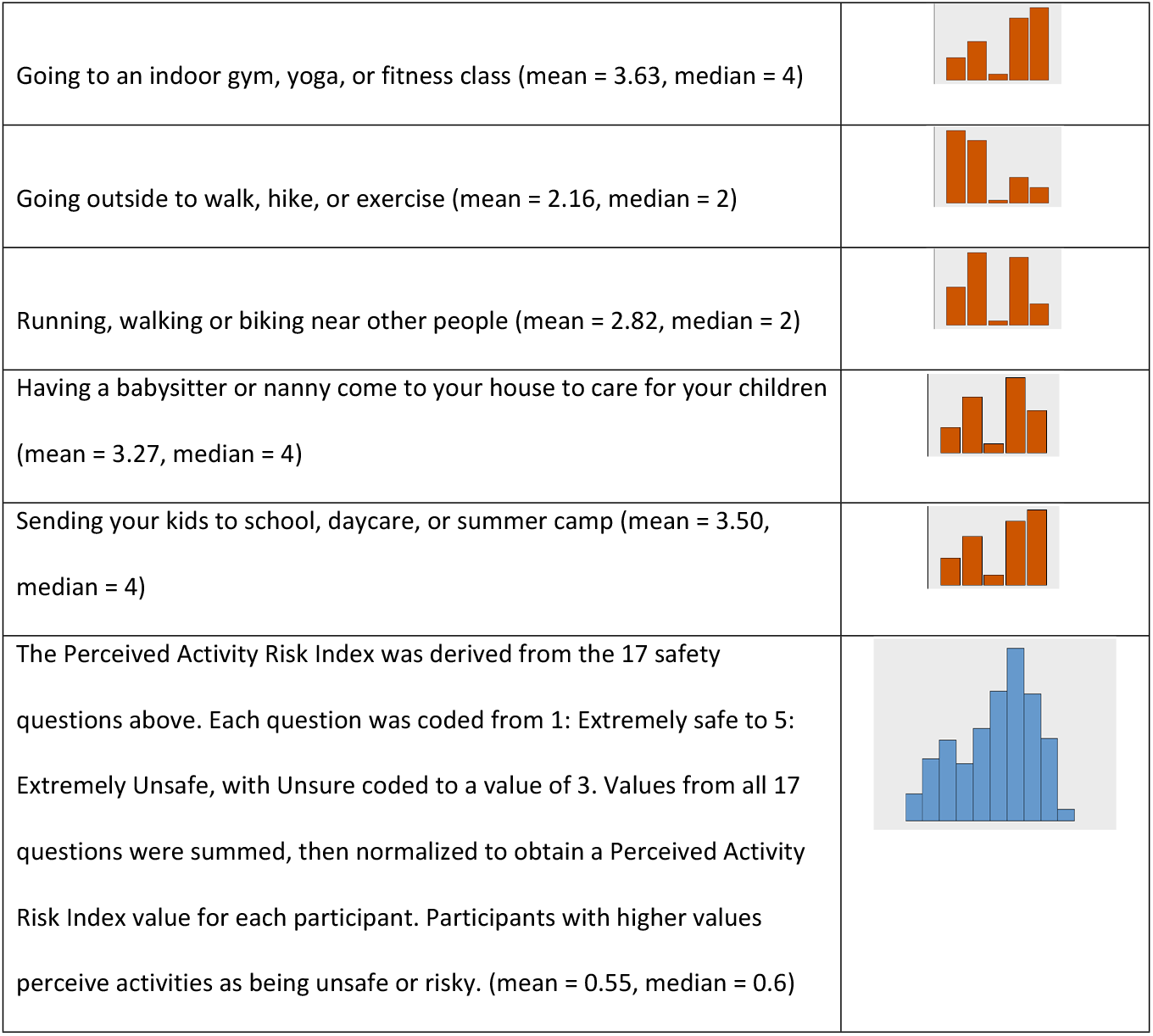
Survey Question: How safe or unsafe are the following actions for avoiding exposure to coronavirus? (1: Extremely Safe, 2: Somewhat Safe, 3: Unsure, 4: Somewhat Unsafe, 5: Extremely Unsafe). Response Distributions: Extremely Safe (left) to Extremely Unsafe (right)

### Serious Game Data and Details

After completing the survey, participants were directed to a computer link that launched the serious game. The serious game was developed using the Unity Engine (Unity Technologies, Version 2019.4.3f1). Participants were prompted with an interactive tutorial before gameplay. The tutorial contained dialog boxes which the participant was required to acknowledge before advancing. These tutorial messages explained the mechanics of the game and the decisions required by the player, highlighted important user interface (UI) elements, and demonstrated how to complete a round of play. After completing the tutorial, participants were given the choice of replaying the tutorial or beginning the experiment. A demonstration version of the game can be found here: https://segs.w3.uvm.edu/CovidBeta

### Serious Game Design and Details

After completing a survey, participants observed an introductory tutorial that describes how to use the game. It is explained to the participant that exposure time in the game reflects more time in the real world: “In this experiment COVID-19 exposure is elevated to reflect the short time that you will be playing the game; seconds in the game represent minutes or hours of exposure. Please consider every second spent near others, within a 6’ radius distance, a meaningful contact with a chance of getting infected from COVID-19.” Moreover, they were told that their performance in the game, the more points they earn, will translate to more U.S. dollars earned. During the tutorial, participants practiced each option in the game setting for obtaining food and obtaining exercise. After completing the tutorial, users completed fifteen rounds each with differing context and messages regarding COVID-19. Alteration of context and messages were considered treatments. Treatments experienced by participants included varying the density of computer-generated people (bots) in game locations, and the infection prevalence among the computer-generated people (infection prevalence information was provided to participants using a visual threat gauge). Treatments specifics are detailed below.

Each round of the serious game was comprised of six scenes, which iteratively follow based on the participant’s decisions. To start each round, scene 1 provided the user with COVID-19 risk information. The second experimental scene prompted the participant to select how they wish to obtain food, by either selecting take-out delivery or going to the grocery store (e.g., Fig. 2). They were informed that take-out will require them to wait for 15 seconds and provides limited points (50 points) or they may decide to go to the grocery store for the opportunity to earn up to 200 points. Going to the grocery store carried the risk of acquiring COVID-19 either through proximity (being within 6-feet of another individual) or through airborne transmission. The longer the participant was in the simulated grocery store environment with potentially infected individuals, the higher their chances of acquiring COVID-19. Participants going to the grocery store were given 30 seconds to collect eight food items, each worth 25 points, for a total of 200 possible points (Fig. 2). After completing food acquisition, the next scene prompted a decision on how to obtain exercise for the day. Participants chose to either go to a park or to exercise at home. Similar to the grocery store decision, the stay-at-home option provides no risk but a low point reward. If participants decide to go to the park, they were tasked with collecting leaves (symbolizing getting exercise in a natural setting) within a 30 second time limit. Ten leaves, each valued at 20 points, were available for collection, allowing for a maximum of 200 total points. In the park environment, participants could become infected if they were within 6 feet of an infected individual. There was no airborne disease vector in the simulated park environment.

**Figure 2.**
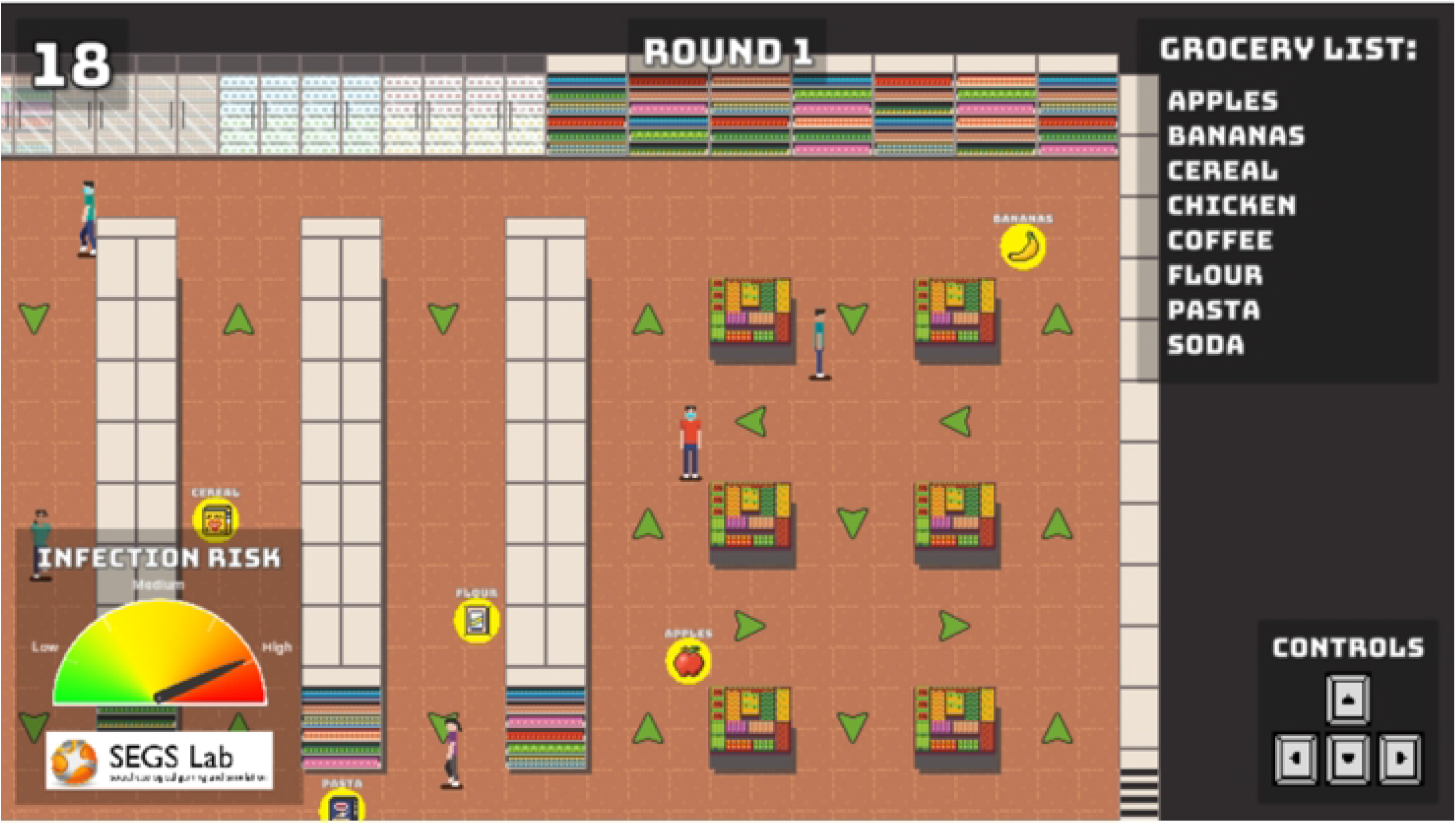
A screen grab of the grocery store environment during the experiment. During the round the participant (center of screen in red) moves around the store potentially interacting with other customers, some of which are wearing masks (e.g., upper left corner) while others are not (blue shirt, center right). The grocery list is displayed in the upper right side. The infection risk is displayed in the bottom left corner with a threat gauge. Time remaining to obtain groceries is displayed in the upper left corner.

After both food provisioning and exercise activities were completed, our interface informed the participant if they became infected, and if so where (grocery store or park) and how much money they earned or lost (in case of infection) during the round. The game then progresses to the next treatment round or an end of game screen if all fifteen rounds have been completed.

In the grocery store and the park environments, participants encountered automated, computer-controlled characters (or “bots”) that navigated around the scene. The number of bots seen in each location was defined by the “density” treatment variable. In the grocery store, bots navigated to different points in the store, pausing at those points for a short amount of time to simulate the collection of groceries before moving on, and eventually leaving the store. In the park, bots were randomly assigned a direction to walk along a trail in the park, and continuously moved in that direction at varying movement speeds relative to the player’s movement speed.

Bots were assigned two possible personalities: risk averse and risk tolerant. Risk averse bots always wore masks and tried to avoid the player in the park by stepping off to the side of the path when the player entered their proximity. Risk tolerant bots never wore masks and did not try to avoid the player in the park.

Every bot was potentially infectious with COVID-19. The proportion of infective and non-infective bots was defined by the *Infection Prevalence* treatment variable, which was communicated to the player using a graphical infection risk threat gauge (Fig. 3). The threat gauge was presented to the participant before each round and additionally was fixed to the bottom left of the simulation interface while navigating each environment. Each bot had a circular “infection zone” measuring 6 feet in radius. For every 0.1 second that the participant was within an infected bot’s infection zone, there was a small chance of infecting the player. Specifically, for every 0.1 seconds within the 6-foot radius, there was a 0.5% chance if the infected bot was unmasked, and a 0.025% chance if the infected bot was masked. In addition to transmission from close contact, we included the potential for airborne transmission in the grocery store. Specifically, to account for increased risk of COVID-19 infection indoors, for every 0.1 second in the grocery store, infected bots in the grocery store had a 0.01% chance (unmasked) or 0.005% chance (masked) of infecting the player, even if the player was outside the bot’s infection zone.

**Figure 3:**
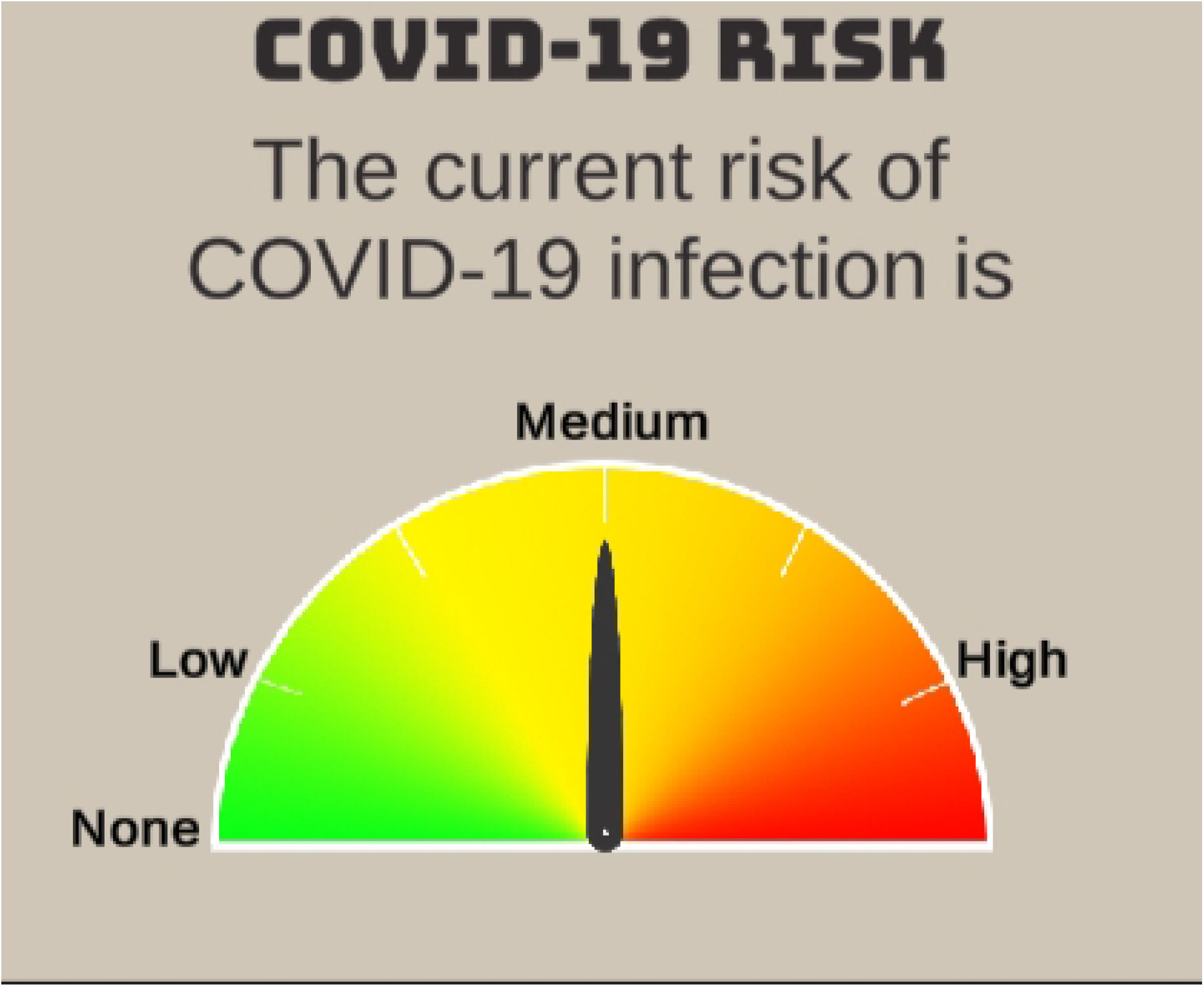
Infection prevalence or risk was provided to the participant before each treatment round and while in the interactive grocery store and park environments.

After completing the round (both food and exercise components, players are informed if they became infected during the round. Infected players receive the message: “You have been infected with COVID-19. You lose 800 points. You were infected at the [Grocery Store or Park]”. After this they see a second screen with the message: “You have recovered from COVID-19. You are no longer infected with COVID-19 and can’t spread the virus. However, you are <u>not</u> immune, and you <u>can</u> become infected again.” The game then proceeds to the start of the next round or to the end of game screen if all rounds were completed.

Throughout the game, we collected data from participants’ choices and actions. Each round, we logged the participants’ decision to go to the grocery store or obtain food using the grocery delivery mechanism; their decision to go to the park or completing a home workout; the time they spent at the grocery store and park; how many groceries and leaves (the unit of exercise quality measurement used in the park environment) they collected; the cumulative time they spent within 6 feet of computer-generated bots; whether they were infected that round; and the points earned. At the end of each round, data were transmitted to a server and stored in a MySQL database.

In summary, during the serious game phase, participants completed fifteen rounds of decision making with four potential decision sets per round. Decision sets were as follows: 1) Deciding how to obtain food: Delivery or Grocery Store. 2) If the Grocery Store option was selected, decision actions while the participant was obtaining food at the store. 3) Deciding how to obtain exercise: At home or going to the park. 4) If the park option was selected, decision actions while the participant was at the park.

### Independent Variables and Experimental Treatments

#### Perceived Susceptibility (Survey)

As previously noted, the Perceived Activity Risk Index, was designed as a measure of perceived susceptibility from the HBM. As such we hypothesized that those with high perceived susceptibility would be more likely to avoid risky situations.

#### Infection Prevalence

The community risk of infection was provided to the participant with the “The risk of infection in your local area is indicated by the risk level dial. Higher risk indicates a higher percentage of infected people in your community.” The infection prevalence or infection risk gauge (Fig. 3) was presented to the participant before each static decision point (food provisioning and exercise provisioning). Additionally, if the participant went to the grocery store or park, the gauge was presented during the entirety of their time spent in those environments. The COVID-19 Risk threat gauge was a measure of the prevalence of COVID-19 in the community.

#### Density of Computer-Generated Bots

Both the grocery store and park environments were populated by computer-generated bots that mimicked the behavior of people in those environments. For example, bots in the grocery store were programmed to collect their own set of grocery store items and move around the store. Density of bots varied by treatment because we hypothesized that the density of people in an environment would influence behavior. Density varied from none (no people in grocery store or park), to low (4 people in grocery store, 8 people in park), to high (8 people in grocery store, 16 people in park). Density was correlated with risk because infection transmission was coded to occur on a probabilistic bases from infected to susceptible individuals.

#### Rounds since last infection (Fear extinction)

The extinction phase of classical conditioning and extinction suggests that removal of stimulus will gradually reduce the behavior associated with the stimulus. In the case of Pavlov’s dogs, salivating after hearing the bell declined until extinction if food was not provided. However, if food was provided during the extinction phase, then salivation returned upon bell ringing to a high level. Similarly, fear behavior can be conditioned (e.g., repeatedly being told that an activity could result in death), and further fear can decay if experience suggests that the fear is unwarranted – called fear extinction learning (38). Here we suggest that the perception of the fear of COVID-19 infection associated with a behavior wanes if no infection occurs. We hypothesize that fear extinction learning may surface in two ways: 1) Fear extinction of grocery store behaviors, and 2) Fear extinction associated with exercise at the park activities. Thus, two variables were quantified to account for potential fear extinction learning: *Rounds since last infection: Grocery Store* and *Rounds since last infection: Park*. These variables measure the length in rounds or experience since an infection: 1) specifically in the grocery store, or 2) specifically in the park. The Rounds since last infection variables are quantified to start at 1 and increased by 1 each round that the participant does not get COVID-19. If the participant does get COVID-19 at either the grocery store or the park, then the appropriate variable is reset to 1 and the process restarts. Thus, a low Rounds since last infection value is associated with low extinction (fear of the activity remains), and a high Rounds since last infection suggests that fear associated with activities may have decreased. Rounds since last infection variables are context specific with the location-specific variables only incrementing upon visiting those locals. For example, the grocery store variable increments up only if the participant goes shopping and does not get sick at the grocery store. It does not increment up if they stay home and have food delivered. The *Rounds since last infection: grocery store* variable resets to 1 if the participant gets sick while shopping. Similarly, the *Rounds since last infection: Park* variable only increments up if the participant goes to the park and does not get sick at the park and resets to 1 if the participant gets sick at the park.

#### Round Number (Learning during Game Play)

The order that treatment scenarios are played was randomized. Yet, players may learn different strategies as the game progresses, such as becoming less willing to go to the grocery store over time because they feel that it isn’t worth the risk. To control for within-experiment learning or behavioral trends we included a covariate labeled *Round Number* (39, 40). To quantify Round Number, we normalized the order that treatments were played from 0 (first round) to 1 (last round), and used this covariate to test if participants changed their behavior as the game progressed.

### Data Analysis

All analyses were completed using R (41) with plots developed using ggplot2 (42). Models were developed for two decision sets: 1) Willingness to go to the grocery store, 2) willingness to break social distancing rules in the grocery store.

#### Willingness to go to the grocery store

We used a mixed effect logistic regression to examine willingness to go to the grocery store to purchase food. The dependent variable was the binary decision to either go to the grocery store or to order food delivery. The model examined included participant ID as the random variable, and the independent variables: 1) COVID-19 community Infection Risk, 2) Rounds since last infection: Grocery Store, 3) Rounds since last infection: Park, 4) the Perceived Activity Risk Index and 5) an interaction term between the Perceived Activity Risk Index and the Infection Risk. No other information was available to the participant when they were making the initial decision to go to the grocery store (e.g., density of shoppers in the store).

#### Aggregate willingness to break social distancing rules in the grocery store

In the simulated grocery store environment participants were required to move using the arrow keys to collect grocery items. Points were accrued by collecting groceries from their list, and infection (resulting in a points cost) could occur from either coming within a 6-foot radius of the computer-generated bots or through simulated airborne transmission.

Of interest was how participant’s perception of the safety of activities for acquiring COVID-19 impacted their willingness to break the 6-foot social distancing rule. We quantified social distance rule breaking across all treatments, including contexts where one was unwilling to go to the grocery store (i.e., they chose to safely procure food through home delivery), avoiding the opportunity to break the social distancing rules. To test the relationship between perceptions of safety and social distance rule breaking, we examined the relationship between the Perceived Activity Risk Index and the total amount of time spent within a 6-foot radius of the computer bots summed across all treatments using a simple linear regression model with aggregate time spent around computer-bots as the dependent variable regressed against the Perceived Activity Risk Index.

## Results

During the serious game phase, participants completed 15 treatment rounds of decision making with four potential decision sets in each round. Decision sets were as follows: 1) Deciding how to obtain food: Delivery or Grocery Store. 2) If the Grocery Store option was selected, decision actions while the participant was obtaining food at the store. 3) Deciding how to obtain exercise: At home workout or going to the park. 4) If the going to the park option was selected, decision actions while the participant was at the park.

The Perceived Activity Risk Index was found to be left-skewed with a mean of 0.545 and a median of 0.603, indicating that participants leaned towards a perception that activities queried were somewhat unsafe. Note the relative polarization in the perceptions of safety with most people not selecting unsure in any of the seventeen individual questions. That is, there are relatively strong beliefs about the safety of activities.

### Willingness to go to the Grocery Store

Results from the mixed-effect logistic regression strongly suggest that the virtual possibility of a COVID-19 infection and the resultant costs influenced participants’ willingness to go to the grocery store with many opting to reduce the amount of money that they took home to avoid the risk of infection. The independent variables considered were the Infection Risk, the Round Order, Rounds since last Infection: Grocery Store, Rounds since last Infection: Park, the Perceived Activity Risk Index (i.e., Table 1), and an interaction term between the Perceived Activity Risk Index and the Infection Risk. We observed strong odds ratios for many of the relationships in the model with reductions in the likelihood of going to the grocery store associated with Round Number (as experience accrued individuals were less likely to risk going to the grocery store), High Infection Risk, and interactions between High and Medium Infection risk messages and the Perceived Activity Risk Index (Table 2, Fig. 4). As described by Figure 4, participants that perceived activities as being generally safe were much more likely to go to the grocery store in both of the higher risk categories than those that described activities as unsafe. However, if the infection risk was Low then those that described activities as unsafe were more likely to go to the store, possibly indicating that those individuals were seeking a relatively safe time to obtain food (and earn consequent money). Figure 5 describes the relationship between the Rounds since last Infection: Grocery Store variable and the Infection Risk. Specifically, in all infection risk situations, as the number of rounds since a participant has become increases, they are increasing likely to go to the grocery store to obtain food. This effect is dramatic in the High Infection Risk treatments, where inexperienced participants (early rounds) or those that have experienced an infection are unlikely to go to the grocery store (i.e., less than 25% choose to go to the store), whereas those that have experienced ten rounds or more without an infection are more likely than not (greater than 50%) to go to the store, indicating that the fear of an infection has largely dissipated for those participants.

**Table 2.**
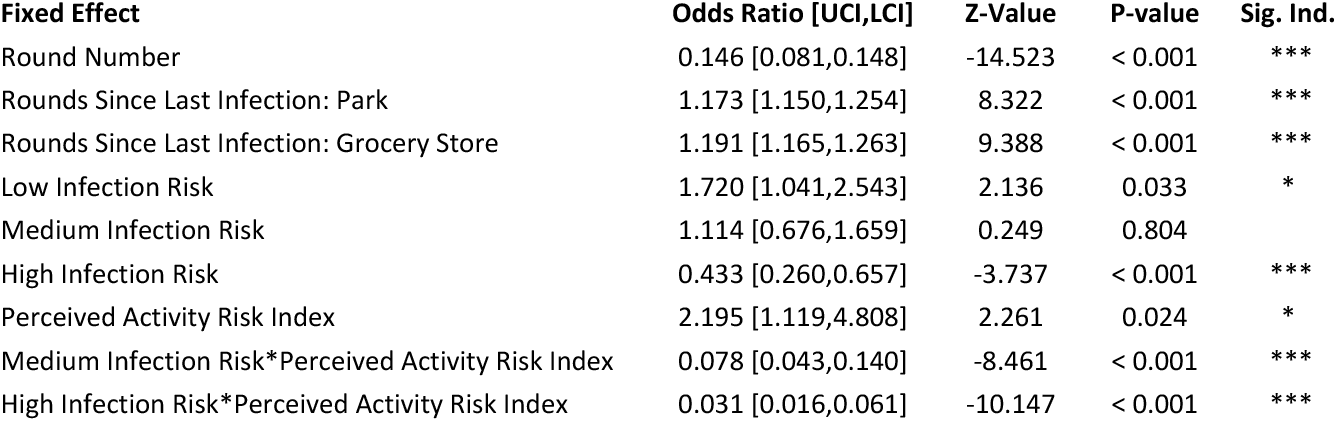
Odds ratios for willingness to go to the grocery store with 95% confidence intervals, Z-values, P-values and indicators of significance.

| Fixed Effect | Odds Ratio [UCI,LCI] | Z-Value | P-value | Sig. Ind. |
| --- | --- | --- | --- | --- |
| Round Number | 0.146 [0.081,0.148] | -14.523 | < 0.001 | *** |
| Rounds Since Last Infection: Park | 1.173 [1.150,1.254] | 8.322 | < 0.001 | *** |
| Rounds Since Last Infection: Grocery Store | 1.191 [1.165,1.263] | 9.388 | < 0.001 | *** |
| Low Infection Risk | 1.720 [1.041,2.543] | 2.136 | 0.033 | * |
| Medium Infection Risk | 1.114 [0.676,1.659] | 0.249 | 0.804 |  |
| High Infection Risk | 0.433 [0.260,0.657] | -3.737 | < 0.001 | *** |
| Perceived Activity Risk Index | 2.195 [1.119,4.808] | 2.261 | 0.024 | * |
| Medium Infection Risk*Perceived Activity Risk Index | 0.078 [0.043,0.140] | -8.461 | < 0.001 | *** |
| High Infection Risk*Perceived Activity Risk Index | 0.031 [0.016,0.061] | -10.147 | < 0.001 | *** |

**Figure 4.**
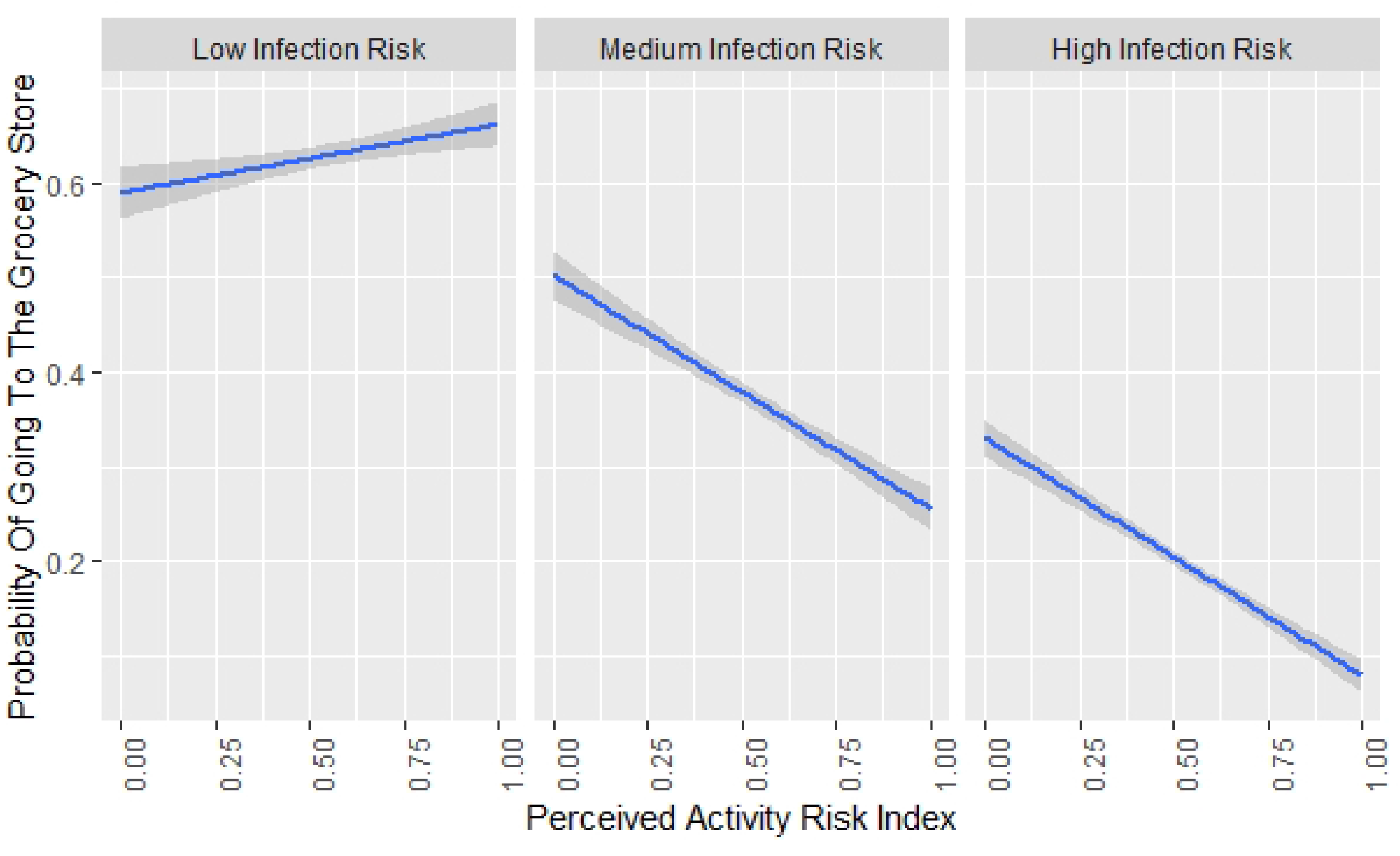
Results find strong contextual relationships between stated preferences and beliefs, and behavior during game play. Infection risk context-specific, willingness to go grocery shopping in the game was correlated with survey responses describing stated perception of the safety of activities such as gathering in numbers. Results indicate that responses to serious game treatments reflect responses of risk typologies to incentives, policies and messages.

**Figure 5.**
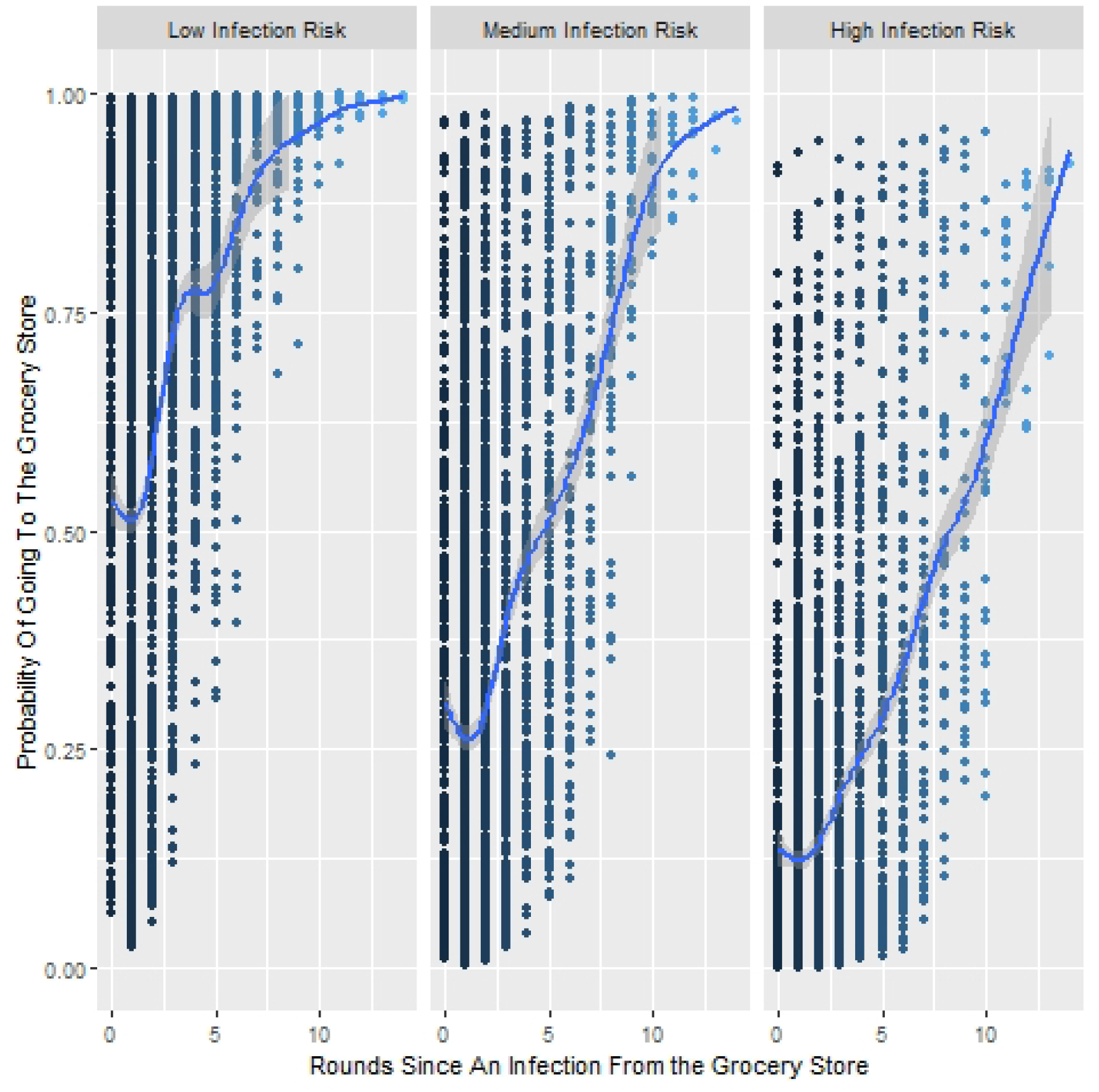
The probability that a participant would go to the grocery store to procure food has a strong relationship with becoming infected at the grocery store on previous rounds with the effect diminishing over time. This fear extinction effect is modified by the prevalence of infection. On the x-axis, 0 indicates that they were infected in the previous round, whereas high numbers indicate that they have consistently gone to the grocery store without becoming infected.

### Aggregate willingness to break social distancing rules in the grocery store

Overall, those that scored low on the Perceived Activity Risk Index (i.e., described activities as general safe) spent significantly more time within a 6-foot radius of people in the grocery store (Effect Size = - 10.319 seconds, standard error = 1.724, t-value = -5.986, p-value < 0.001) than those that described activities as unsafe (Fig. 6).

**Figure 6.**
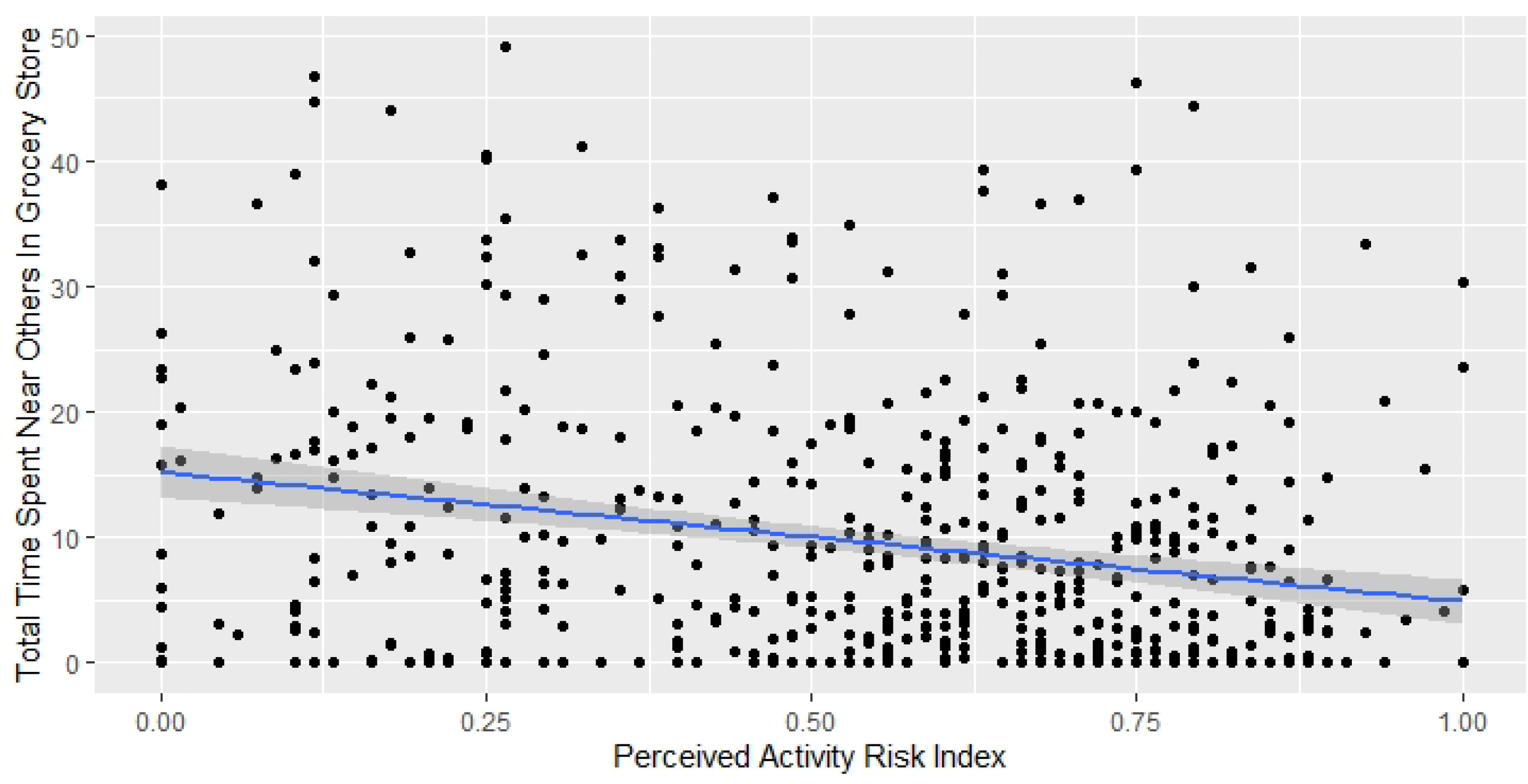
The survey-derived Perceived Activity Risk Index describes perception of how risky a variety of activities are for acquiring a COVID-19 infection. This index was strongly correlated to how much time participants spent near others in the virtual serious game environment with those that felt that activities were generally risky spending significantly less time breaking the social distancing rule than those on the other side of the spectrum.

## Discussion

Mixed-method approaches to obtain data on potential responses to risk messages, communication strategies and novel policies allow for the triangulation of data, thus potentially generating predictions that are more robust than using any single methodology (e.g., surveys, focus groups, or serious games). Moreover, using user-friendly data gathering approaches, such as serious games, promote engagement and quick responses.

We found that infection risk level was a strong predictor of user’s willingness to go to the grocery store, with High infection rates dramatically reducing user’s willingness to go to the store (OR: 0.433). Medium infection risk was also correlated with slightly increased odds of going to the store (OR: 1.114), and a strong willingness to go the store was associated with Low infection risk scenarios (OR: 1.720). Also note the strong interaction effect with the Perceived Activity Risk Index: when the infection risk was medium or high, individuals were much more likely to go to the grocery store if they believed that activities were generally safe, contrasted with those individuals that believed that activities were risky. In the real-world perceptions of susceptibility are likely confounded by other factors such as ideological-driven messaging and cultural norming, which makes the infection risk gauge possibly overly simplistic. However, this also points to the value of providing a simple device, or messaging strategy such as a scaling infection risk indicator.

As participants accrued experience, they went to the grocery store much less frequently, opting to obtain food through the home delivery mechanism (OR 0.146). This indicates that participants were willing to alter their strategy during the process to adjust to the perceived risks and rewards offered by going to the grocery store (i.e., they could obtain four times as many points (200 vs. 50) but risked losing 800 points).

The Rounds since last Infection variables used in the willingness to go to the grocery store model suggested that fear of becoming infected, and possibly the consequent perception of the probability of an infection declined substantially and increasingly with repeated risk-taking and without becoming infected. Specifically, number of rounds since getting infected at the park (OR: 1.173) and number of rounds since getting infected at the grocery store (OR: 1.191) both increased the likelihood that a participant would go to the grocery store. This effect is illustrated in Figure 5 where willingness to go to the grocery store increased the longer and more successfully that participants had avoided becoming sick at the grocery store, with probability modified by the prevalence of COVID-19 in the community (infection risk). With high prevalence of COVID-19, an individual who either avoided taking risks or who recently took risks and became infected was less likely to go to the store than an individual who had taken risks and avoided getting sick for a majority of the game. This relationship was apparent but less pronounced at lower infection risk levels. This pattern indicates that individuals update their risk perceptions of an activity depending on whether they get infected after engaging in that activity. In this scenario, the cognitive process of fear extinction (38), where iteratively engaging in risky behavior without consequence may result in disillusionment or extinction of the fear of infection, and thus may lead individuals to update their risk perceptions in a way that biases their subjective risk perception compared to the objective risks.

The Perceived Activity Risk Index was created from survey data to generalize user perception of how likely or unlikely they were to contract COVID-19 across a selection of activities (Table 1). We found correlations with this index and behavior during serious game play, especially when considered within an infection risk context. When participants were told that the community infection risk was Low, individuals were less likely to go to the grocery store if they believed that in general activities were safe and unlikely to lead to a COVID-19 infection (OR: 2.195) but much more likely to go to the store when infection risk was Medium (OR: 0.078) or High (OR: 0.031) (Fig. 4). This interaction demonstrates a strong correlational relationship between perceptions and in-game behavior. Moreover, that behavior and perceptions interact with the simulated risk context. These results support the HBM-derived hypothesis suggesting that perceived susceptibility to a disease, in this case COVID-19, will impact behavior, with the context (infection risk) and the survey-queried Perceived Activity Risk Index both aligning with perceptions of susceptibility. Therefore, communication strategies that nudge understanding of perceptions of susceptibility should impact behavioral responses. Over the course of game play, those that scored low on the Perceived Activity Risk Index, e.g., described activities as general safe, spent significantly more time within a 6-foot radius of people in the grocery store. Those that were at the extreme lower end of the Perceived Activity Risk Index spent more than three times as much simulation time in close proximity to the computer-generated bots than those on the extreme upper end of the spectrum. Again, this reinforces that perceptions of risk and susceptibility influence behavior even in a simulated environment. We should stress that because this is a simulated environment, perceptions of risk of activities should not change behavioral strategies from a rational actor perspective, yet these results describe correlations between risk perceptions in the real-world and how participants “play” in a simulated environment.

Analyses of the willingness to go to the park and behavior within the park were completed but have not been reported here for purposes of brevity. Overall, the data align well with decisions to go to the grocery store and behavior in the grocery store, i.e., the data support that participants with low Perceived Activity Risk Index scores were more likely to go to the park, and overall spent more time breaking the 6-foot social distancing rule within the park environment. These results provided little additional insight but added complexity. For example, decisions to go to the park for exercise appeared to be influenced by the experience in the grocery store environment, including an index created to quantify time spent breaking the 6-foot social distancing rule, including whether they obtained food by food delivery. A successful grocery store experience (quantified as going to the grocery store and having less than average time breaking the social distancing rule per treatment) was positively correlated with going to the park to obtain exercise. Overall, differences between the grocery store and park environments were not radically different, perhaps because at the time of data collection, it was less recognized that outdoor interactions were unlikely to lead to significant COVID-19 exposure.

Because participation in this serious game was completed before wide-spread availability of vaccines, perceptions formed regarding the safety of non-pharmaceutical interventions such as social distancing were likely related to behavior in real world situations. The ability to see these relationships reflected in game play, with risk perceptions (i.e., the Perceived Activity Risk Index) strongly predicting the likelihood of engaging in risky behavior during the game, provides evidence that we can delineate individuals that may be more likely to take risks in everyday activities, and further, identify regions with subcultures that may have a generally consistent set of risk perceptions, that may promote or dampen the spread of disease. Identifying these subcultures, through queries about the safety of activities, may allow for targeted messages that nudge perceptions of susceptibility, and thus nudge behavior to more conservative, risk mitigating behaviors.

Our research shows that survey responses to perceptions of the risk associated with activities correlated to their risk behaviors in the simulated environments. This finding, though not surprising, demonstrates the efficacy of cognitive theories that assert that perceptions shape actions (e.g., HBM or the Theory of Planned Behavior (43)), even in contexts such as simulations or online games that, according to rational actor theory, should be distant from actual behavior. That is, the HBM and Theory of Planned behavior might suggest that perceptions are more tightly linked to real-world behavioral intentions than hypothetical behavioral intentions.

These findings also shed light on the important role that public health information plays in governing behavior, suggesting that risk information does, indeed, impact behaviors of people who hold beliefs across the spectrum of perceptions about the safety or riskiness of activities. Those with a high degree of risk safety are most sensitive to information and thus may be more responsive to frequent and fluid risk communication strategies. This suggests that public health information campaigns and infographics such as real time risk and threat indicators, should be widely used to alert the public to elevated or lowered risks and opportunities to shape perceptions of risk situations are possible and likely needed. These findings also demonstrate the efficacy of using serious games as proxies for behavior - games which can leverage theoretical underpinnings such as the HBM to further our understanding of decision-making and behavior. The capacity of serious games to provide dynamically changing environments or contexts, vary treatments in information and risk rates, and observe responses generates more nuanced data that can be leveraged to understand within-subject variability. These data coupled with perceptions elicited via survey data, provide a powerful mixed methods approach to studying human behavior. Considerations such as participant socio-economic background, political ideology, and geographic locations can provide subtle, but important differences in the efficacy of risk communication strategies. In addition, these findings provide finer grain understandings of participants’ willingness to adopt which can allow for more geographically bound profiles of sub-populations, allowing public health officials to tailor messaging and policies to trigger behavioral nudges. Future work could delve into additional social and cultural factors, such as ideologies or geographical factors, that may be linked to the perception of susceptibility or the perception of the benefits of using COVID-19 risk mitigating behaviors. Further work could identify vaccine-hesitant individuals and use a virtual, serious game to determine which policies, social environments and communication strategies alter their behavior in a virtual world, which could provide insight into how to effectively increase vaccine uptake.

## Conclusion

Here we show that a user-friendly data gathering tools, serious games combined with surveys, could be leveraged to generate big data, with potential for understanding and testing interventions to improve risk communication, policy design with resultant public health implications. Results explore the relationship between perceptions, intentions and simulated behavior – a critical step in linking intent to behavior.

Overall results depict strong relationships between stated beliefs and observed responses to experimental conditions or contexts. This suggests that messages, policies and communication strategies can be tailored to stated belief structures and assessed for efficacy. Consequently, we can quantify the relative effects of strategies and policies before deployment and reduce the likelihood of perverse incentives or effects, and further, suggest best communication strategies to achieve intervention goals.

We show that behavior of individuals within a virtual environment could be used to help understand a participant’s belief structure about the perception of activities in a COVID-19 environment, and vise-versa. It follows that the virtual environment can then be used to test messaging and communication strategies to determine their effectiveness for nudging behavior, or nudging correlates of behavior such as perceptions of susceptibility. Thus, we can explore messages and policies to determine efficacy for nudging behavior of people that hold different sets of beliefs, allowing for the targeting and optimization of strategies to reduce risk-taking behavior and lessen the impact of COVID-19.

## Data Availability

Data will be held in a public repository and available after acceptance.

## Acknowledgments

We acknowledge support from the Social Ecological Gaming and Simulation laboratory at the University of Vermont.

## Notes

### Competing Interest Statement

The authors have declared no competing interest.

### Funding Statement

CJK, SCM, AZ, TS Internal grant from the University of Vermont's Gund Institute for Environment, Burlington, VT. USA (https://www.uvm.edu/gund). SAN, JPH Internal grant from the University of Vermont's Translational Global Infectious Diseases Research Center, Burlington, VT. USA (https://www.med.uvm.edu/tgircobre/home). The funders had no role in study design, data collection and analysis, decision to publish, or preparation of the manuscript.

### Author Declarations

The University of Vermont’s Institutional Review Board approved this research Approval number# STUDY00001027. Titled: COVID-19 Outbreak Behavior Experimental Simulation Game and Survey. Consent was obtained using online using computer, and was considered a type of written consent.

